# A process evaluation of a cluster randomised trial hosted in hairdressing salons promoting women’s cardiovascular prevention

**DOI:** 10.64898/2026.03.01.26345507

**Authors:** Jennifer Y. Barraclough, Menglu Ouyang, Michelle Reading, Mark Woodward, Anthony Rodgers, David Peiris, Anushka Patel, Bruce Neal, Clare Arnott, Hueiming Liu

## Abstract

**Aim:** To outline the opportunities and barriers when using hairdressing salons as a novel site for enhancing cardiovascular risk factor assessment and management in women.

**Methods:** A process evaluation nested within a cluster-randomised trial, ‘Hairdressers for Health’. The trial evaluated a ‘nudge’ intervention advising women ≥45years attending hairdressing salons to undertake a ‘Heart Health Check’ with their General Practitioner. The UK Medical Research Council process evaluation framework was used to guide the design, data collection and analysis. Nineteen interviews were conducted with nine hairdressers, nine study participants and a project officer. Thematic analysis assessed recruitment, reach, acceptability, and adoption. Characteristics of the salons and participants were analysed using descriptive statistics.

**Results:** Recruitment of the planned 88 metropolitan and 28 regional salons for the trial was challenging, requiring resource-intensive face-to-face visits. The nudge intervention was well accepted by participants, and salons were perceived to be an appropriate setting to effectively reach women. Adoption of the study by salons was limited with only 54 of the 116 salons recruiting participants (total recruited 239, range 1-22 participants per salon). Barriers to participant recruitment included technological constraints while using a decentralised online recruitment and data collection platform, client preferences and privacy concerns. Established hairdresser-client relationships in smaller salons facilitated greater client participation and was perceived as a good mechanism for health promotion.

**Conclusions:** Cardiovascular health prevention messaging for women in salons was acceptable to hairdressers and clients. Designing the study to make better use of hairdresser-client personal relationships may have improved project implementation.

**Trial Registration:** ACTRN12621001740886

## Introduction

Cardiovascular (CV) disease is a leading cause of death in adults globally, resulting in 18 million deaths (1) and is responsible for the death of 22 women every day in Australia. Australian women have high rates of poorly controlled cardiovascular risk factors, with over 60% of the female population overweight or obese and in those over 45years of age more than 60% having dyslipidaemia. In addition, 20% of Australian women have uncontrolled hypertension (2). As a result, the National Heart Foundation of Australia have advocated for a primary healthcare “Heart Health Check” (HHC), to address risk factors and thus prevent cardiovascular events. The HHC is a Medicare Benefits Schedule (MBS) item number, funded by the government, allowing General Practitioners (GPs) to be reimbursed for performing a formal cardiovascular risk assessment on a patient over the age of 44 years. However, only 1.1% of the 1.6 million eligible women ≥45 years in NSW, Australia, accessed a HHC in 2020(3) and, in the 2 years leading up to 2018, national data suggested only 39% of Australian women aged ≥45 years had any sort of heart risk assessment performed by a health professional (4).

Non-healthcare-based community settings for health intervention have been shown to be effective in providing health messages to promote awareness, including in beauty salons(5) and barber shops (6–8). The “Barber Shop study,” undertaken in the USA (7) successfully engaged African American males with hypertension in treatment, from barber shops. This was a pragmatic clinical trial designed to evaluate the effectiveness of an intervention in hairdressing salons, which aimed at accounting for real world variables in adherence, uptake and practice among healthcare providers(9).

The Hairdressers for Health Study was designed as a pragmatic cluster randomised trial using a “nudge” intervention, conducted in hairdressing salons in New South Wales, Australia. The intervention aimed to encourage women 45 years of age and older to undertake a HHC with their GP, and the study aimed to understand the potential for promoting cardiovascular risk assessment outside traditional healthcare settings. This setting was specifically chosen as 52% of Australian women ≥50 years visit a hairdresser over a 4-week period.

Pragmatic trial design aims to recruit participants who are similar to patients who would receive the intervention if it became usual care and measure outcomes relevant to ‘real-world’ environments. It bridges the gap between research evidence and practice that is directly applicable to real-world healthcare decision-making. Since pragmatic trial design uses less resources, they come with several challenges such as variability in implementation, participant heterogeneity and variable data quality including missing data(10). In addition, there are often challenges relating to stakeholders not having prior research experience and lack of sufficient research infrastructure to handle trial work(11). In pragmatic trial designs, the intervention should be delivered as in normal practice. Blinding may not be feasible or consciously avoided to prevent impact on the intervention; however, allocation concealment should always be implemented(12).

Process evaluation from implementation trials provides information on the challenges and feasibility of implementation of the intervention and translation of findings (13) to improve healthcare provision. Several pragmatic trial domains were used in this trial including participant eligibility criteria, intervention flexibility, follow-up intensity and participant adherence to study protocol(14). It is valuable in unpacking for whom, how and why an intervention had an impact on the study outcomes, as well as providing insights to guide future trial design. The aim of this process evaluation was to understand the recruitment, acceptability, adoption and barriers to using hairdressing salons as a novel site for enhancing cardiovascular risk prevention in women, to inform future pragmatic trials in non-healthcare settings.

## Methods

### Trial design and context

The Hairdressers for Health Study, a cluster randomised trial, was hosted in 120 hairdressing salons in New South Wales. Salons were randomised 1:1 to intervention and control arms at the time of recruitment of salons and both the salons and researchers were blinded to the intervention allocation. Salon location and sociodemographic region was measured by Socioeconomic Indices for Areas (SEIFA)(15). 27 participants from 120 salons were planned to be recruited, providing 80% power to detect 5% absolute difference in the primary outcome between groups, assuming 2 alpha = 0.05 and ICC of 0.1. The participant was introduced to the study through a flyer with a QR code in the salon, which they scanned with their personal device, which took them to a bespoke electronic study platform. The participant was advised to read the participant information form and provide informed consent to participate, at which point they were directed to the online health questionnaire. In the intervention arm participants were given a CV health pamphlet and advised to attend their GP to undertake a HHC, with a dedicated letter explaining the study to take to the GP (**Supplementary Appendix 1**). The control arm was provided with a general women’s health pamphlet only (**Supplementary Appendix 2**). The salons and study staff were blinded to intervention allocation. Thereafter, data linkage through MBS and PBS (Pharmaceutical Benefits Scheme) and a follow-up survey was undertaken at 6 months (**Figure 1**). The study was approved by human ethics committees Australian Institute of Health and Welfare # EO2021/5/131 and University of New South Wales # HC210621.

**Figure 1:**
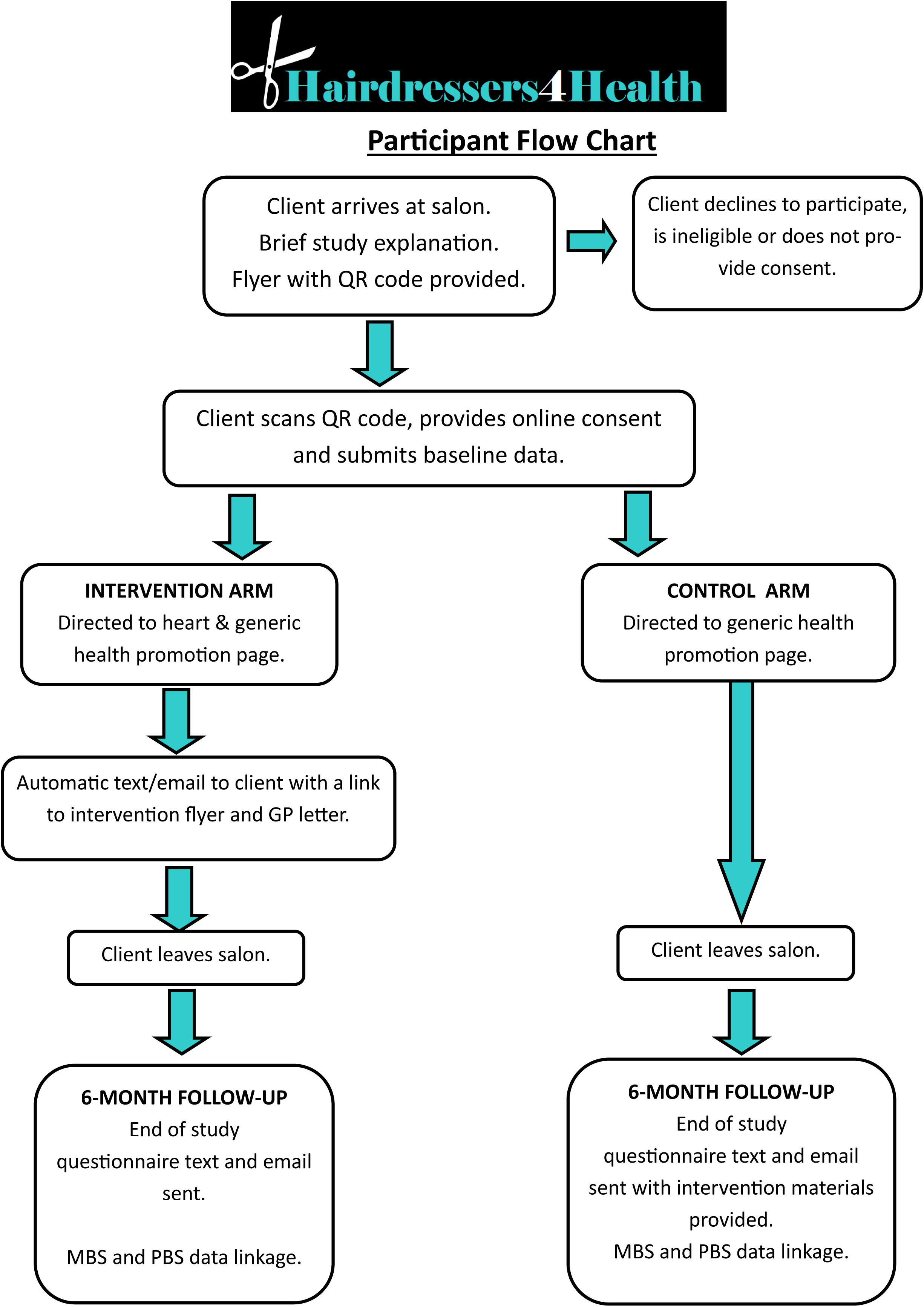
Study Flow Chart Process of Clinical Trial. Study flow chart description of the cluster randomised trial process. GP (general practitioner), MBS (medicare benefit schedule), PBS (pharmaceutical benefit scheme).

### Trial Data Collection

The bespoke study platform allowed participants to directly enter their demographic data, baseline questionnaire and 6-month follow-up questionnaire answers, online. Participants accessed the website at 6 months via an embedded link in an email/text message sent by the study team.

Baseline data included mandatory demographic and contact information including name, date of birth, contact email address, home address, mobile phone number and the baseline health survey was undertaken by participants during the salon visit or at a later time. The Trial Baseline Health Questionnaire Data (all self-reported) can be found in **Supplementary Appendix 3**.

The Follow-up 6-month Health Questionnaire administered via the study website (or telephone) with additional questions regarding whether the study impacted on their interaction with the healthcare system, can be found in **Supplementary Appendix 3.** This facilitated participant selection for interview prior to delayed linked data health outcomes. Data Linkage was carried out for all participants using PBS and MBS datasets held by the AIHW to assess the use of the HHC MBS item numbers at 6 months. Data were accessed and protected through the Centre for Health Record Linkage (CheReL).

### Process evaluation methodology

The process evaluation was informed by the UK Medical Research Council process evaluation guidance(16) and the implementation research logic model (17) to unpack the contextual determinants and mechanism of implementation outcomes of the intervention. The process evaluation was undertaken post-trial to understand the intervention implementation and the insights into barriers and facilitators for the pragmatic trial that was conducted in a non-healthcare setting. The intervention, contextual determinants, implementation strategies and theorised mechanisms are summarised in **Figure 2**.

**Figure 2.**
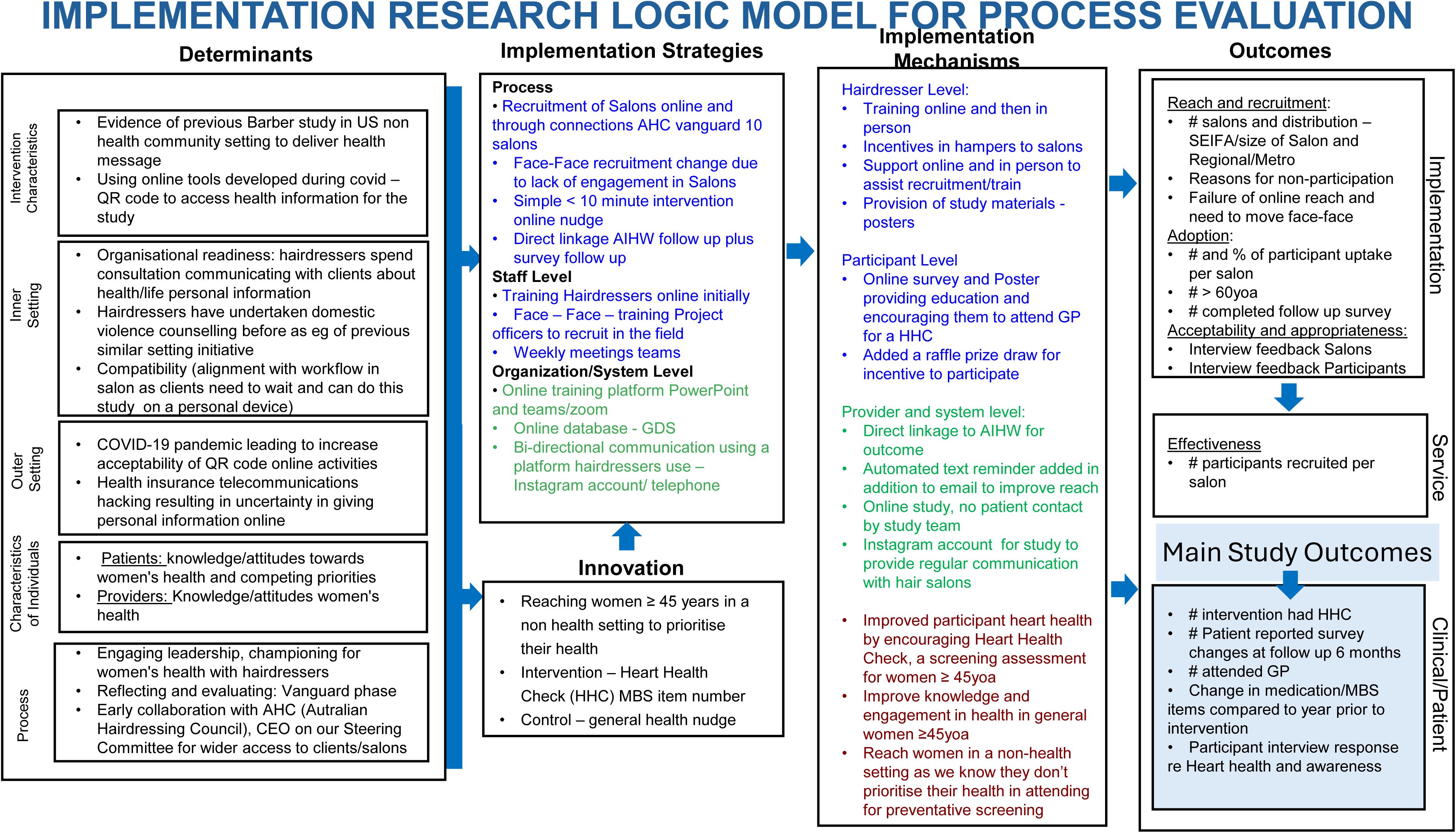
Implementation Research Logic Model for Process Evaluation. Main study outcomes listed in the bottom right represent pending data for the main study awaiting data linkage with Australian Institute of Health and Welfare. GDS (Data Management System); MBS (Medicare benefits Schedule)

### Process Evaluation Qualitative Data

The data collection materials were piloted in the trial vanguard phase to finalise the interview guide (**Supplementary appendix 4**) and optimise trial implementation processes. The post-trial process evaluation interviews were conducted with hair salon staff, participants and a project officer between 21st Oct 2024 to 22nd November 2024 to gain insights from diverse stakeholders on the study conduct and implementation through the approach of grounded theory(18). All interviews were conducted post study closure of recruitment, and 6 month follow up of participants. A convenience sampling approach was used to select the participants based on their availability and willingness to participate. Three study team members (MR, KN and SH) were trained by experienced experts in qualitative research (HL and MO) to conduct interviews. All interviews were conducted online or by telephone and audio recorded and transcribed verbatim without transcripts being returned to participants for comments or correction. There were 24 salon staff approached to take part in the interview by telephone and email, and 9 completed the interviews. Among 57 participants who were approached for interview, only 11 agreed to participate and the interviews were completed in 9. Reason for non-participation included “too busy” or no response on contact. In total, 19 interviews were completed for the process evaluation (9 with salons, 9 with participants and 1 with the project team). The characteristics of the participants are shown in **Table 2 and 3**. There was no prior relationship to the participants with the interviewer apart from the project officer who worked on the project with the research team.

**Figure 3.**
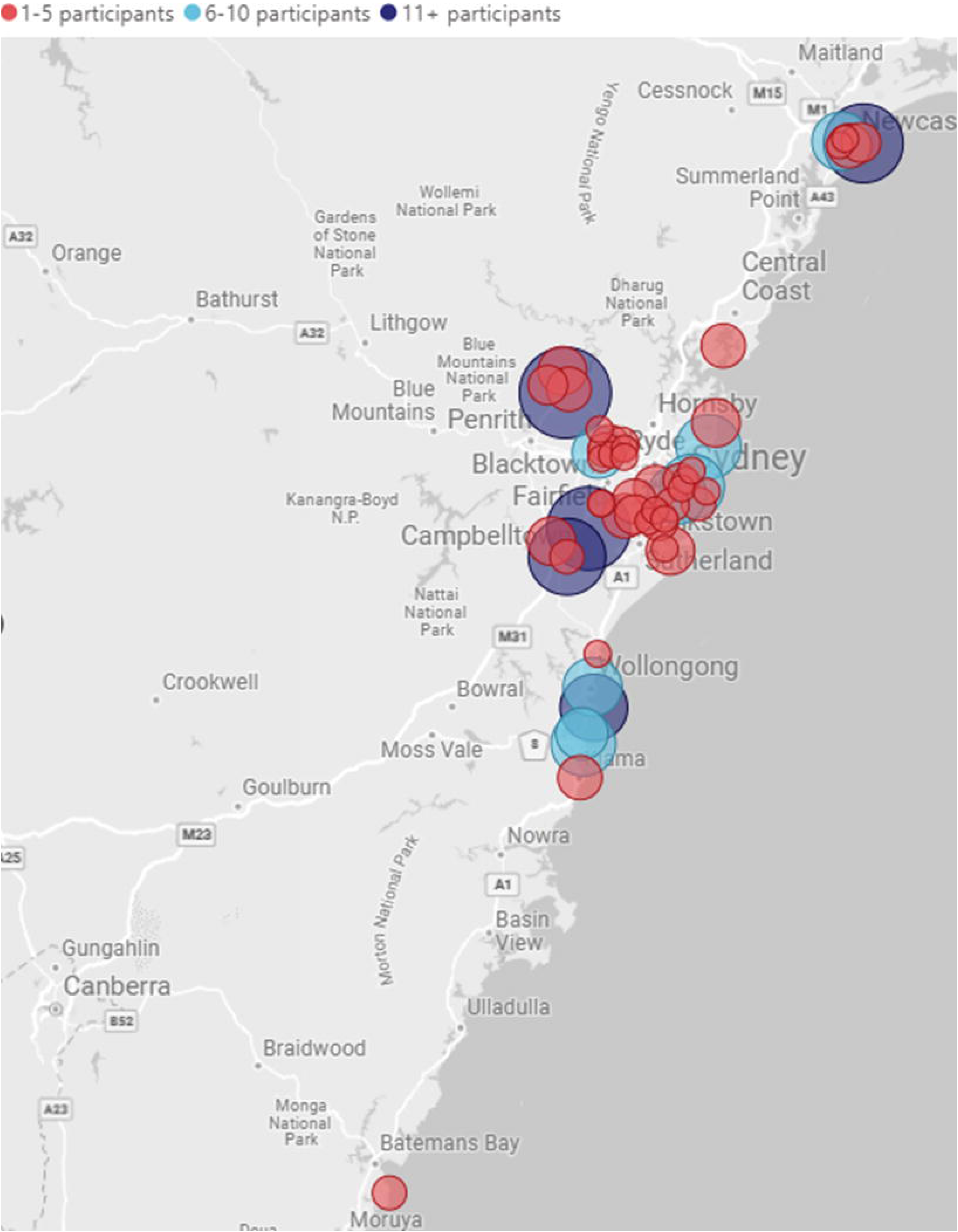
Location Map of Salons by Number of Participants Recruited. Salon location throughout New South Wales in Australia. Each circle marker represents a salon in the study, and the size and colour represent the number of participants recruited. 1-5 red and smallest circle, 6-10 light blue and intermediate sized circle and 11 or more participants dark blue and largest circle.

### Process Evaluation Quantitative Data

The demographic information of the salons and participants was collected from the baseline questionnaire. Salon data included SEIFA score, metro or regional location, total participant enrolment and proportion over 60 years. In addition, salons were categorised into low (0–5), intermediate (6–10) and high recruiters (11 or greater) (**Figure 3 and Table 1**). Study participant data included demographics such as age, education level, non-English-speaking background, Aboriginal and Torres Strait Islander and % completion of baseline and survey at 6 months (**Table 2**).

**Table 1.** Study Salon Characteristics.

| **Salon Characteristics** | **Control (N)** | **Intervention (N)** |
| --- | --- | --- |
| Salons enrolled | 59 | 57 |
| Salons Actively recruiting | 27 | 27 |
| No. Regional Salons | 12 | 16 |
| No. Metro Salons | 47 | 41 |
| **SEIFA** |  |  |
| 1 | 11 | 12 |
| 2 | 10 | 11 |
| 3 | 10 | 8 |
| 4 | 10 | 9 |
| 5 | 18 | 17 |
| **High recruiters > 10** | 6 | 2 |
| **SEIFA by high recruiters** |  |  |
| 1 | 3 | 1 |
| 2 | 0 | 0 |
| 3 | 1 | 0 |
| 4 | 0 | 1 |
| 5 | 2 | 0 |

“Intervention” includes salons providing Heart Health Check specific information and “Control” involves salons providing a general health nudge. SEIFA is the socioeconomic indices for areas (15) N = whole number.

**Table 2.**
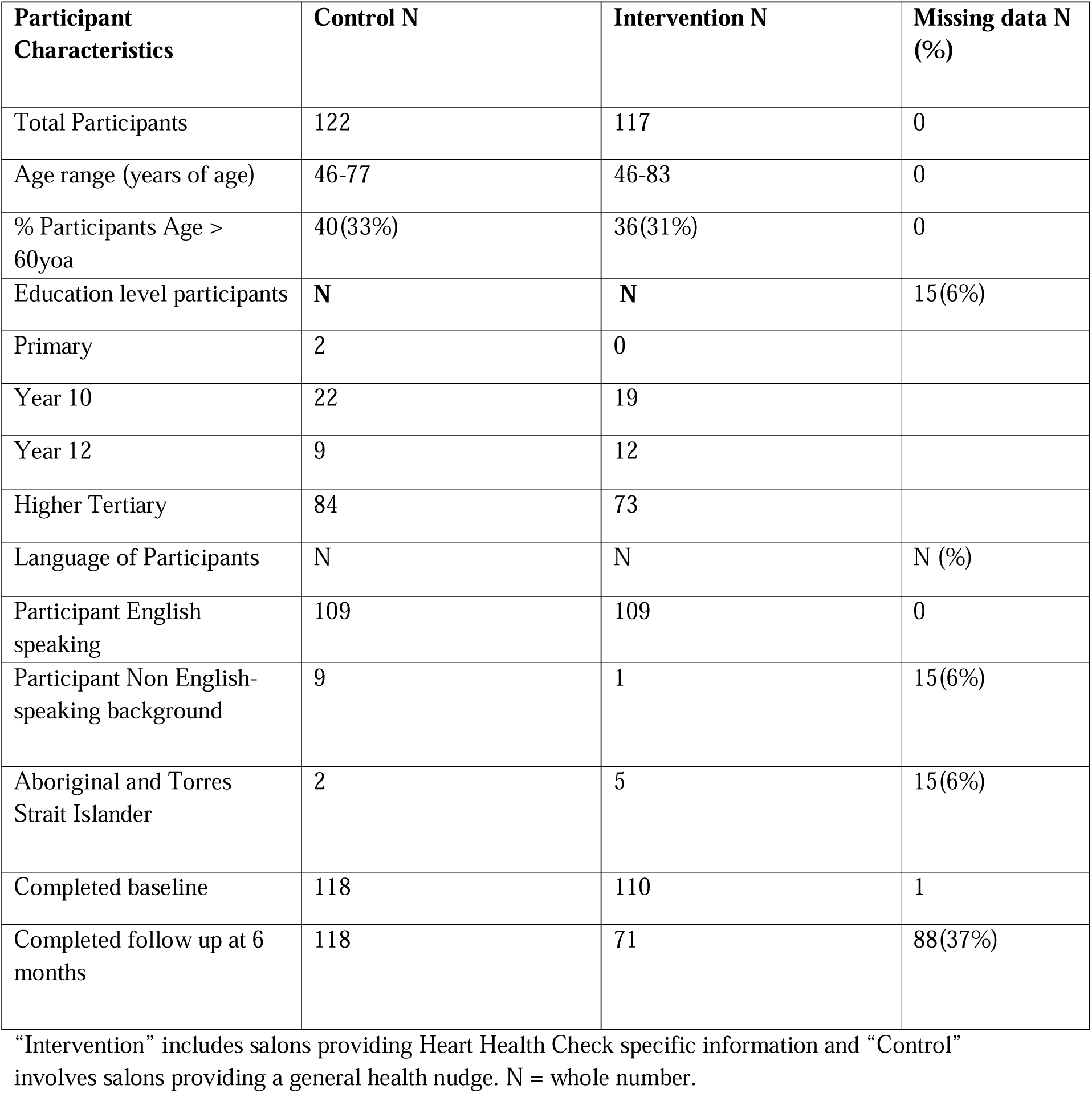
Participant Characteristics.

### Data analysis and reporting

Participants data were analysed using descriptive statistics, on Microsoft Excel (version 2506) and Microsoft Power BI (version 2.145.1105.), and reported in numbers and percentages for reach and adoption. For qualitative data, thematic analysis using inductive coding was conducted based on the implementation research logic model (**Figure 2**), to assess contextual determinants and the mechanisms that result in the implementation outcomes of reach, acceptability, appropriateness, and adoption of the intervention. Audio recordings were transcribed to de-identifiable qualitative data and analysed. A codebook was developed using an inductive coding of the first 5 interviews by MO. The themes were discussed with the study team (MR and HL) until the thematic saturation was reached with no new codes created. Nvivo 11.0 was used to manage the data and the SRQR checklist standards for reporting qualitative research were used to report the findings from the interviews (19). Findings were triangulated using the implementation research logic model (**Figure 2**)

## Results

### Reach and Recruitment

Reach was challenging as it depended on the recruitment of hairdressing salons which were new to research, and hairdressers recruiting the relevant participants based on their eligibility criteria. The study recruited 116 salons across metropolitan and regional areas (of a target 120) though only 54 (47%) of them actively recruited participants to the study, ranging from 1-22 participants per salon (**Figure 3 and4**). The characteristics of the salons, such as location and SEIFA, were balanced between the intervention and control groups (**Table 1 and 3).** However, there were more high recruiter salons in the control group than in the intervention group (6 vs. 2). The barriers and facilitators to optimise the study recruitment and implementation are summarised in **Table 4**. There were 88 Salons from metropolitan areas and 28 from regional areas in NSW, with active recruitment of participants from 1-13 months and an average participant recruitment speed of 2/month with a change in recruitment strategy with face-to-face recruitment, which increased the number of participating salons within a short period of time (**Figure 4**).

**Figure 4.**
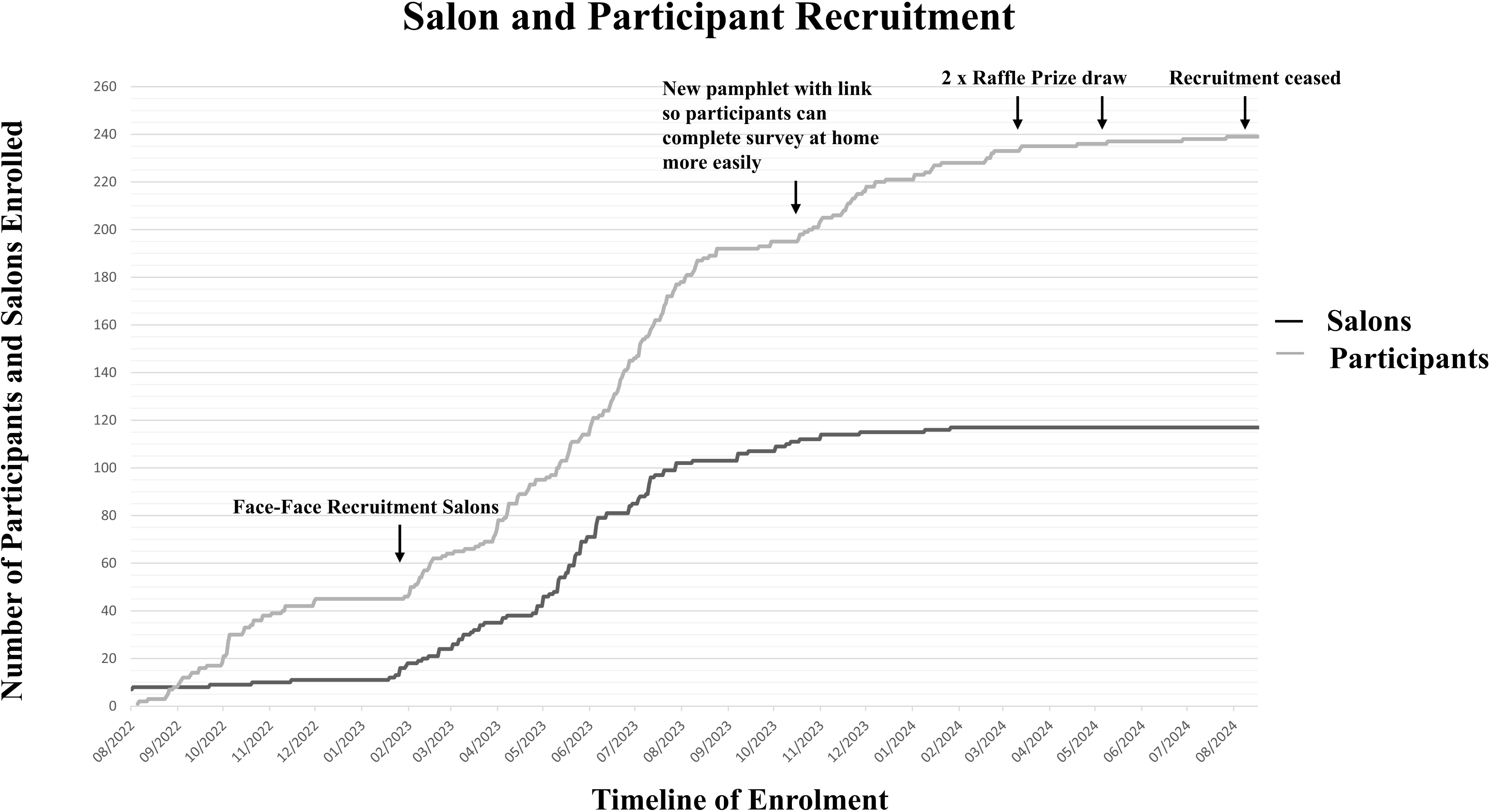
Recruitment Strategy and Trajectory of Salons and Participants. Line graph representing timeline of recruitment and trajectory for both Salons and Participants recruited to the study. Time points of interventions aimed to boost recruitment. X axis represents months from study commencement. Y axis represents whole number of salons and participants recruited.

**Table 3.**
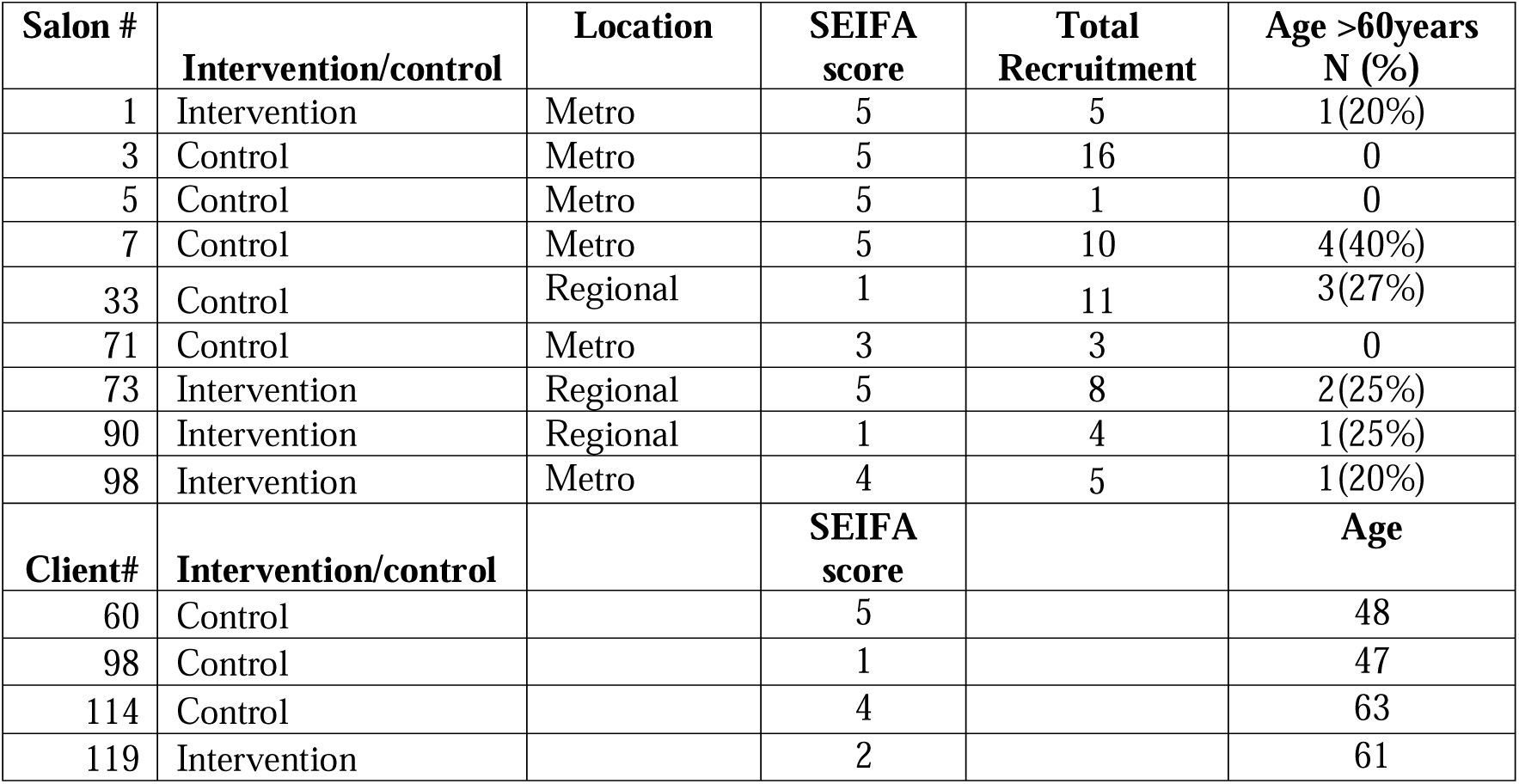
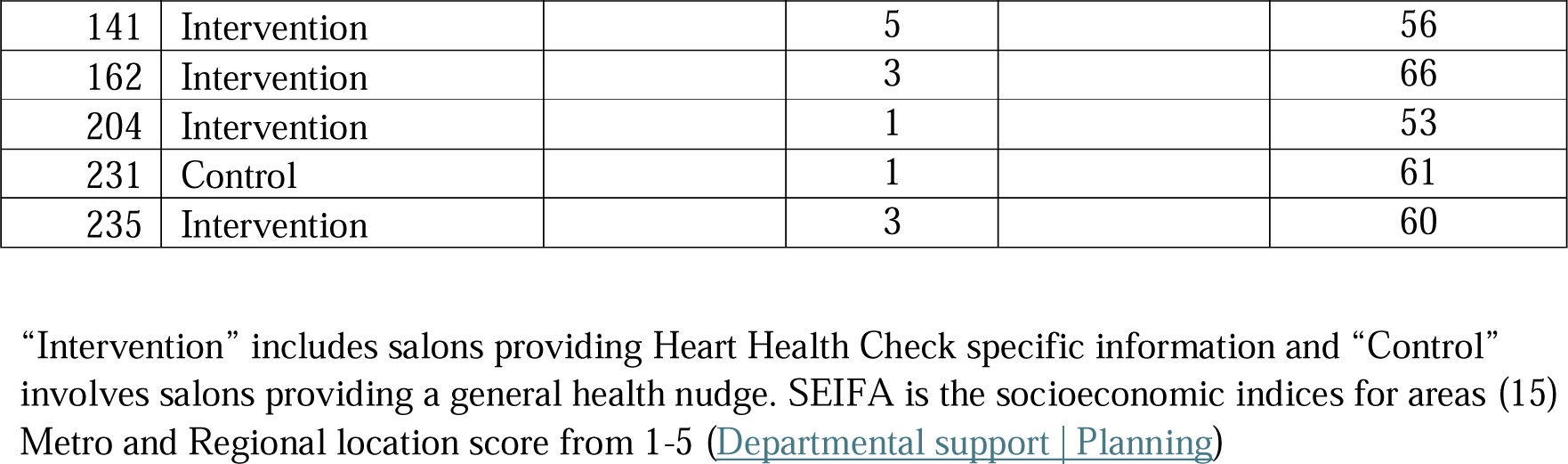
Characteristics of the participating Salons and Clients in the Process Evaluation.

| **Salon #** | **Intervention/control** | **Location** | **SEIFA score** | **Total Recruitment** | **Age >60years N (%)** |
| --- | --- | --- | --- | --- | --- |
| 1 | Intervention | Metro | 5 | 5 | 1(20%) |
| 3 | Control | Metro | 5 | 16 | 0 |
| 5 | Control | Metro | 5 | 1 | 0 |
| 7 | Control | Metro | 5 | 10 | 4(40%) |
| 33 | Control | Regional | 1 | 11 | 3(27%) |
| 71 | Control | Metro | 3 | 3 | 0 |
| 73 | Intervention | Regional | 5 | 8 | 2(25%) |
| 90 | Intervention | Regional | 1 | 4 | 1(25%) |
| 98 | Intervention | Metro | 4 | 5 | 1(20%) |
| **Client#** | **Intervention/control** |  | **SEIFA score** |  | **Age** |
| 60 | Control |  | 5 |  | 48 |
| 98 | Control |  | 1 |  | 47 |
| 114 | Control |  | 4 |  | 63 |
| 119 | Intervention |  | 2 |  | 61 |

|  |  |  |  |  |  |
| --- | --- | --- | --- | --- | --- |
| 141 | Intervention |  | 5 |  | 56 |
| 162 | Intervention |  | 3 |  | 66 |
| 204 | Intervention |  | 1 |  | 53 |
| 231 | Control |  | 1 |  | 61 |
| 235 | Intervention |  | 3 |  | 60 |

“Intervention” includes salons providing Heart Health Check specific information and “Control” involves salons providing a general health nudge. SEIFA is the socioeconomic indices for areas (15) Metro and Regional location score from 1-5 (Departmental support | Planning)

**Table 4.** Barriers and facilitators with example quotes.

|  | Illustrative themes | Impact on | Example quotes |
| --- | --- | --- | --- |
| **Barriers** | Internet/ technical issues | Study conduction | *“Among all, I think. But she didn't have a good Internet at her salon, for example, even my phone wasn't working there. So that was a big issue, I think.” (Project officer)*<br>*“That sounds completely fine and normal, but then obviously the practical side of doing it is slightly different, because then you're left to that your own devices and you don't always know exactly what you're getting them to sign up to do” (Salon #5)*<br>*“-OK, so you didn't scan a QR code in the hairdressers to complete a survey now (interviewer).*<br>*-No, you're talking to someone that hates QR code. (interviewee)” (Client #114)* |
|  | Busy workload of hairdressers | Recruitment | *“Probably just that we kept forgetting. You just forget about it yet, because you just forget to hand it out.” (Salon #73)*<br>*“Overlook a lot of things because of our busy schedules. So I think something that makes someone stop and sit for a minute and read it and go.” (Salon #33)* |
|  | Privacy concerns | Recruitment | *“And they (clients) were asked if they had the Medicare like, why do you want my Medicare number or, I don't know, if ask the number actually, but that was actually a question. If they have the Medicare and then they were like I don't, I don't think I want to do that.” (Project officer)*<br>*“I think the biggest thing that went wrong for you guys as a whole was just all the personal information that people had to fill out. I had a lot of people start to fill it out and then was like, no, I'm not feeling this out any further.” (Salon #33)* |
|  | Client preference | Recruitment | *“With more so like not elderly, but middle-aged, middle-aged women, now it's a younger frontier vibe and you know people are pulling me aside saying I don't like this.” (Salon #71)*<br>*“some of the younger ones it may be a bit daunting, and if there's a 20 year old trying to broach the subject about Women's Health in that sector, they might find it a bit challenging.” (Salon #5)* |
|  | Hairdresser lack of training or unfamiliar with the project | Intervention delivery and recruitment | *“Yes. Yeah, I didn't. Didn't know enough about it (training). Probably my own fault. I didn't really read into anything much 'cause. I don't have. Like, yeah, I have e-mail and all that sort of thing. But for me to get to my e-mail, I don't check my emails through the day and then by the time I get home at the night, that's the last thing I'm thinking about doing is check an e-mail.” (Salon #1)*<br>*“I know that you the envelope is to show people, but I think another thing that would be good is for the person talking about it to understand exactly what the questions are that are going to be being asked because it's alright to ask someone to.” (Salon #5)* |
| **Facilitators** | clients' trust and hairdresser's encouragement | Recruitment | *“I guess it comes from the source, so if it's, if it's given out from someone that you know and that you trust or someone that you know.” (Client #231)*<br>*“No issue with that at all, because I spoke to Lady who came in. She was very informative and I knew that it was a trusted source, so I just tried to reinforce that with people that it was a trusted thing.” (Salon #33)*<br>*“It could be because of their (hairdresser) health issue they had in the past and they wanted to help their clients...some of them are not interested. They prefer to be looking at Instagram or talking or doing other things.” (project officer).* |
|  | Familiarities to the information | Intervention delivery and | *“I think we almost need a bit of a blurb, a bit of a brief of how to explain it. I think when the envelopes were given to* |
|  |  | recruitment | *us, that was.” (Salon #98)*<br>*“it's done online and the paper envelope that I'm giving them cause I didn't wasn't sure what was inside that envelope, but after you explained to me that it's just copies of what they're going to see on the questionnaire. It made it clearer cause um, I thought that there was going to be referrals and all of that kind of stuff in there for more personal information.” (Salon #5)* |
|  | Appealing promotion materials | Recruitment | *“If it was modified so that the shorter and easier to pitch, and then for the close to then be done by other people other than the hairdressers that are trying to do hair.” (Salon #90)*<br>*“Definitely wanted to speak about that. Um, maybe the posters they got changed twice, but maybe the posters could be more eye catching. Maybe a little bit, maybe a bit broader or something” (Salon #33)* |
|  | Study team engagement | Intervention delivery | *“But. Even hairdressers are better support skill set so that they can deal with that information is probably something that will need to be done if health services are going to work more this way to engage with.” (Salon #5)* |
|  | Study setting | Recruitment | *“They are small salons have regular clients and they don't have the new clients...but big salons have the names they have like they in this case, they don't have a regular Clients so it keeps rotating so they don't have the good relationship with their clients.” (Project officer)*<br>*“I think a hairdressing environment can be. It's sort of like the cone of silence that where people feeling comfortable in discussing different things and also being able to take whatever information is given to them, be it a pamphlet or whatever, and feel.” (Client #114)* |

The client journey in a hairdressing salon is depicted in **Figure 1**, highlighting the flexibility in recruitment in a community-based setting. There were several barriers to recruitment and adoption of participants. A total of 239 participants from 54 of the 116 salon sites were recruited by the hairdressers and completed the informed consent. Among the 239 enrolled participants, 32% of them were over 60 yrs (**Table 2**). 230 (96%) completed the baseline questionnaire and 151 (63%) completed the follow up questionnaire at 6 months. Salon characteristics influenced the reach of participants such as the size and location of the salon as *“small salons have regular clients and they don’t have the new clients…but big salons”* may not *“have regular clients”* and thus larger salons may have a greater number of participants to approach (Project Officer, **Table 4**). The recruitment of participants was also driven by the hairdresser’s preference and personal conditions such as *“their (hairdresser) health issue they had in the past and they wanted to help their clients”. (Project Officer,* **Table 4**) Inner setting barriers were also mentioned by hairdressers, such as having *“good internet at salon” (Project Officer)*, technical difficulties with the online ‘nudge’ intervention and forgetting to provide study information due to the busy workload at the salon (**Table 4**).

### Acceptability: Mechanisms to increase the engagement of women

The “nudge” intervention increased engagement of women. The study aim was well accepted by salon staff and participants, to approach women in hairdressing salons to improve CV risk factor identification and disease prevention using an online pamphlet and questionnaire. The salon staff stated that it is a good idea to reach women in salons: *“I actually really liked it. I thought it was really a clever way to. I just feel like it’s a place where mostly women hang out and in my salon particularly, we have group conversations and stuff all the time about all the topic that we’re told. I’m pretty sure that’s about Women’s Health and like, when did you have your last medical and stuff like that and you don’t have to answer it to me. But I think it’s really important to put that information out there to help others.” (Salon #33)* Hairdressers described how discussion of health themes and lifestyle factors are common in hairdressing salons and therefore acceptable. From participants, a hairdressing salon was an acceptable setting to attain information regarding women’s health as it provided an environment “*where people feeling comfortable in discussing different things”* and take on information (Participant #114).

### Adoption of the trial in non-healthcare setting

Participant’s characteristics, such as age and background, influenced the decision to sign up and complete online questionnaires. The trial was designed to utilise developments in information technology to conduct the study with personal devices with a QR code. Hairdressers described that some participants had significant concerns about privacy in entering personal information to consent and to complete long questionnaires, in the context of data privacy and security. “*I think the biggest thing that went wrong for you guys as a whole was just all the personal information that people had to fill out. I had a lot of people start to fill it out and then was like, no, I’m not feeling this out any further.”* (Salon #33, **Table 4**)

Hairdressers reported barriers in the salon workflow in the study set up, with hairdressers forgetting to provide the paper envelopes, consisting of the paper version of the online pamphlet in addition to the online version*. “Probably just that we kept forgetting. You just forget about it yet, because you just forget to hand it out.” (#Salon 73)*

The salon staff reported not being aware if clients signed up or not and were uninformed of progress in recruiting their target of 27 women per salon. This feedback led to the researchers implementing a strategy of informing salons of progress in recruitment of participant numbers by their salon. However, there was still uncertainty of whom had signed up, raising concerns for the hairdressers in approaching existing clients in their salon more than once, which could negatively affect the hairdresser-client relationship (**Figure 5**).

**Figure 5.**
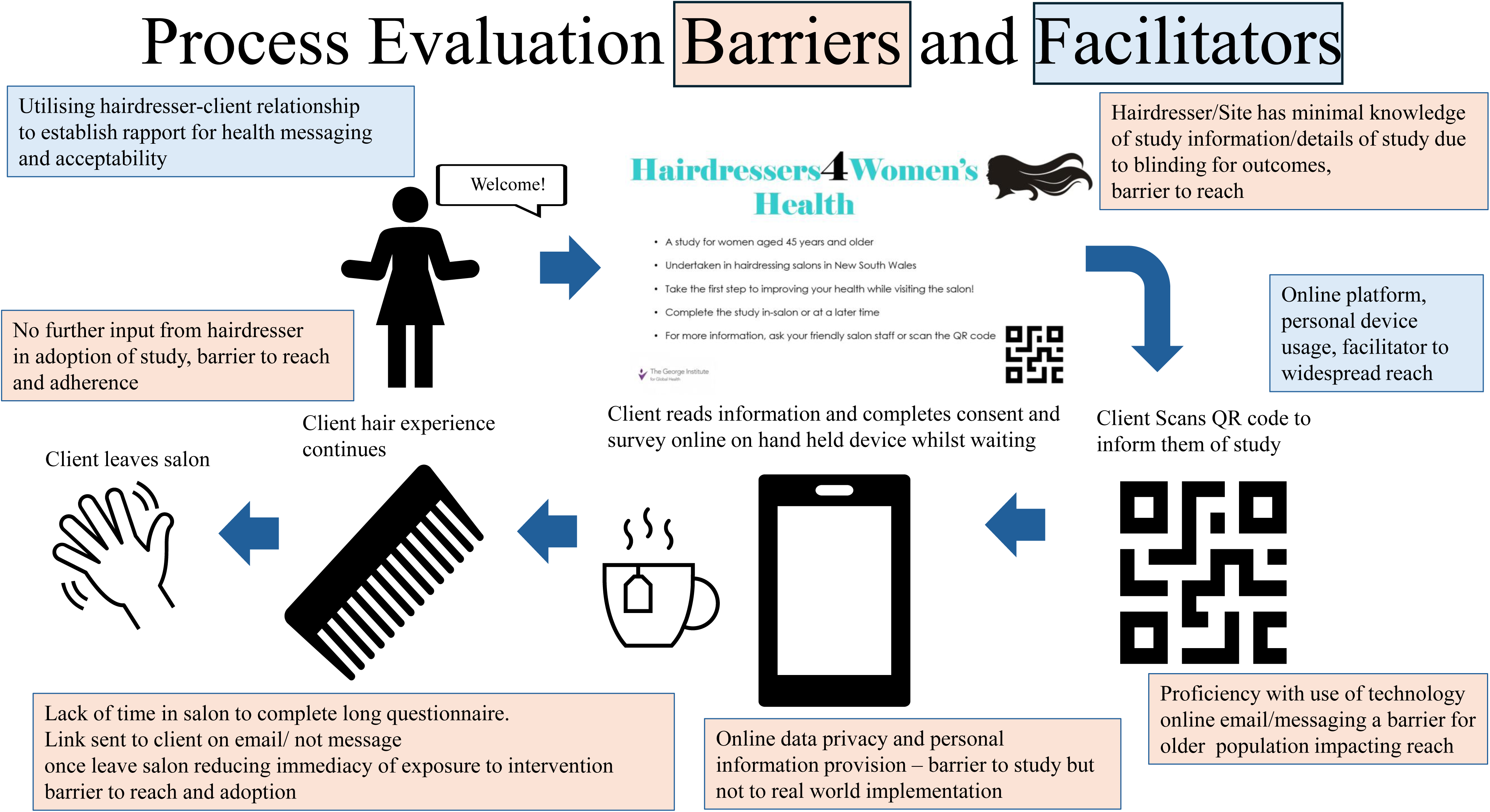
Process Evaluation Barriers and Facilitators. Graphical representation of the barriers and facilitators this process evaluation study identified as contributors to the adoption, recruitment and adherence for the cluster randomised trial.

Participants recalled having received the flyers or information on the study, however, some of them reported not following up with seeing their GP to undertake a HHC. Adoption of the study may have been reduced due to the decentralised recruitment and lack of follow up contact after the “nudge” intervention: *“The hairdressers are on board with it then yes, because if they forget to talk to their clients about it, they’re just the flyers and the posters are just going to just sit there…It was quite a long time ago and I signed up for it, but then I didn’t do anything else after that.” (Participant#204)*. Minimal study follow-up online through email and text reminder only, as a pragmatic ‘nudge’ intervention trial design, was aimed to target behaviour in a low cost, low resource manner. Hairdressers suggested that participants should be reminded to take action after signing up to the study, to improve the uptake of the health information package. It was suggested that further interactions between the study team and clients would, improve adoption of the intervention by the participants. *“Got some clients with QR code first and then you guys can contact them…And then I can hand over the rest of the project to you to make sure that they sign it because there’s no point.”* (Salon #90).

### Facilitators to Recruitment and Adoption of Participants

Hairdresser-client relationships and the importance of the clients’ trust, and encouragement from the hairdressers, was deemed to be a key element for the acceptability of women’s health promotion in a hairdressing salon. The hairdressers stated more information and familiarity with the health package would improve their confidence to talk to their clients. *“I think we almost need a bit of a blurb, a bit of a brief of how to explain it. I think when the envelopes were given to us, that was.” (Salon #98), “ I think another thing that would be good is for the person talking about it to understand exactly what the questions are that are going to be being asked because it’s alright to ask someone to.” (Salon #5)* Therefore, the blinded trial design, may have reduced the reach to participants.

Streamlined training was provided to the salon staff which included a brief explanation, provision of materials to view and an online training session. Although online, in person and telephone support was offered to salons throughout the study by researchers, this mostly was not taken up by salons as it was not a priority for the hairdressers, as illustrated by a hairdresser’s comment (Salon #1) “*I don’t check my emails through the day and then by the time I get home at the night, that’s the last thing I’m thinking about doing is check an e-mail.” (Salon #1)* Suggestions included establishing a closer relationship with the salon staff at the beginning of recruitment and/or participants, to improve study familiarity, understanding and research confidence amongst salon staff may have improved the recruitment of participants. Other suggestions included developing more appealing materials and a personalised approach to the health-related materials provided to their clients (**Figure 5**).

## Discussion

The Process Evaluation for this study identified several barriers and facilitators to the conduct of a pragmatic cardiovascular trial in women within hairdressing salons. The setting of this clinical trial represented a community-based environment and non-healthcare setting frequented by women, which could opportunistically engage women in hairdressing salons to undertake a HHC. Active recruitment and utilising established relationships with hairdressers to facilitate trial implementation is pertinent to success of reach of the trial in this community-based setting. Similarly, adoption in salons and by clients is attributed significantly to the personal relationships and follow up by the hairdressers who have established rapport with the clients. Prioritising use of these hairdresser-client relationships will heavily influence adoption by salons and participants.

Several aspects of the pragmatic trial design influenced trial success. The decentralised trial design, with online recruitment of individuals at the hairdresser, using a QR code on a personal device, meant minimal follow up and contact with the individuals for trial adoption and adherence, to assess the intervention in a real-world environment with email and text reminder only. This may have contributed to loss of follow-up and study adoption and recruitment, though we used linked data for our primary outcome using MBS and secondary outcomes data through linked PBS outcomes and survey follow-up aimed to minimise measurement bias.

### Active vs Passive Recruitment

The recruitment of participants in this trial was passive, requiring the participant to scan the QR code, and sign up for the trial and then review the pamphlet and act on the information accordingly. This was felt to result in significant reduction in reach and loss to follow up. Although this requires less resources, we reflect that this passive recruitment limited the reach of participants in a research naive setting. Indeed, while passive recruitment is often more cost effective, less resource intensive and pragmatic, it may result in reduced recruitment. (20). Suggestions of more personalised materials used in the salon and co-design with the hairdressers to utilise the hairdresser-client relationship may optimise the recruitment process. Recruitment of hairdressing salons was also initially passive, with promotion of the study through the Australian Hairdressing Council (AHC), relying on hairdressing salons to approach the research team to participate. The change to an active process of recruitment with face-to-face salon recruitment after the Vanguard phase significantly improved recruitment rates. This suggests that active recruitment in a community-based setting is needed, a finding that has similarly been demonstrated in other healthy-lifestyle interventions in the community(21).

### Decentralised data collection

Decentralised trial design, with online recruitment of individuals poses risks to loss of follow up within the trial. Though, the unobtrusive collection of trial outcomes through linked data, is attractive, as it reduces the burden on investigators (22). The online provision of the intervention in this study and text message and email follow up was successful regarding the acceptability and ease of adoption by the salon sites. Evidence suggests a text message can encourage health promotion and is effective in cardiovascular secondary prevention messaging utilising digital resources to improve engagement (23–26) and there is increasing evidence that e-Health intervention improves health outcomes(27, 28), increasing dissemination and navigating geographical and financial barriers and is supported by the WHO.(29). However, the pragmatic ‘nudge’ design may have reduced the reach and adoption of participants as a result of lack of repetitive exposure and trackability of recruitment of participants by salons. Furthermore, the need to input personal identifiable information into the study platform to participate in the study was seen as a barrier to recruitment due to health privacy. When utilising online messaging in primary cardiovascular prevention trials, in community settings, privacy and data protection will greatly influence reach and acceptability and needs to be carefully considered. The barriers experienced in this study, in this regard, however, are not real-world implementation barriers as health messaging in real-world practice would not require the input of personal information to track the study outcomes as was required in this study design.

### Trusted Relationships and health messaging

Previous studies in community-based health interventions have shown “trusted” relationships contribute to participation and efficacy of interventions. This has been shown to be related to personalised care, knowledge translation and individual behaviour (30).

In our study the participant reach was impacted by personalised relationships with a higher proportion of regional hairdressing salons being moderate or high recruiters to the study compared to metropolitan salons. However, smaller salon client pool in regional areas may also result in slower recruitment over time, due to lower turn-over of clients eligible for the study. Personal relationships with illness and a view to helping their community has been shown to contribute to participation and retention (30) evident in our study as hairdressers reflected on being less able to drive participation and trust in the study due to blinding and a lack of information. The unpacking of the Barber Shop study through process evaluation demonstrated the importance of this relationship to the receptivity of participants to health education and leveraging existing relationships (30). This relationship leveraging was limited in our pragmatic trial design due to the blinding of salons and the lack of contact between researchers/GPs and hairdressers to leverage established relationships and develop new relationships with health care providers. The importance of established relationships should thus be considered to deliver effective implementation of a non-healthcare-based intervention.

### Strength and limitations

This study involved diverse participants including clients, hairdressers and the project operation staff to provide insights on a non-clinical setting study. Though a large sample of the participants, were invited to participate, there was a limited number of participants available for interview, and these were not balanced by demographic, and other important characteristics, which might lead to volunteer bias. The timing of conducting the interviews was at the later stage of the trial, and it was challenging for the clients to recall the study process or the materials they received for intervention delivery.

### Conclusions

A non-healthcare-based setting of hairdressing salons is an acceptable setting for cardiovascular health messaging. Reach, adoption and recruitment in a non-healthcare-based cluster randomised study are determined by personal relationships and participant factors which are not uniform and need to be carefully considered to improve implementation.

## Data Availability

All data produced in the present study are available upon reasonable request to the authors

## Abbreviations

CV: cardiovascular
HHC: Heart Health Check
GP: General Practitioner
MBS: Medicare Benefits Schedule
PBS: Pharmaceutical benefits Scheme

## Declarations

### Ethics approval and consent to participate

The study was approved by human ethics committees Australian Institute of Health and Welfare # EO2021/5/131 and University of New South Wales # HC210621.

#### Consent for publication

– Not applicable

#### Availability of data and materials

**-** The datasets used and/or analysed during the current study are available from the corresponding author on reasonable request.

### Authors’ contributions

JB, ML, CA and HL contributed to the design, analysis, interpretation and drafted work, have substantively revised the work and have approved the submitted version.

MR contributed to the design and analysis and drafted work and has approved the submitted version.

MW, AR, DR, AP and BN contributed to the design, interpretation and substantively revised the work for submission and approve the submitted version.

## Acknowledgements

We would like to acknowledge the data management team at the George Institute for Global Health for the development and maintenance of the study online platform. We would also like to acknowledge Samantha Hand and Kellie Nallaiah study project officer and project manager for the Cluster RCT for their contribution to qualitative interviews. We would like to acknowledge the contribution of the CEO from the Australian Hairdressing Council Sandy Chong, for her support and contribution to study design, community engagement and recruitment of hairdressing salons.

## Supplementary Appendix

1. Poster for Intervention arm
2. Poster for Control arm
3. Questionnaire Clinical Trial
4. Interview Guides
5. ISQR Checklist

