## Supplementary Materials for "A process evaluation of a cluster randomised trial hosted in hairdressing salons promoting women’s cardiovascular prevention"

Did you know?

### Heart disease kills

### 22 women every day in Australia

- High blood pressure, high cholesterol, smoking, diabetes and obesity increase the risk of having a heart attack or stroke
- Women who have experienced high blood pressure or diabetes in pregnancy are also at an increased risk of developing heart disease

#### Most women are unaware they are at risk of heart disease:

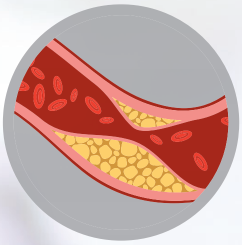

**1 in 3 women** have high cholesterol

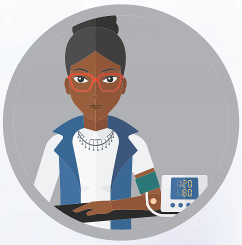

**1 in 5 women** have high blood pressure

#### To assess your risk you should:

- Visit your GP regularly for the following checks:

| | |
|---|---|
| - blood pressure | - cholesterol |
| - weight | - blood sugar |

- Talk to your GP about your heart risk and what you can do to reduce your risk

#### We ask you to:

- Visit your GP within the next month with this letter
- When you book the appointment, please let reception know you will be asking your GP for a Heart Health Check

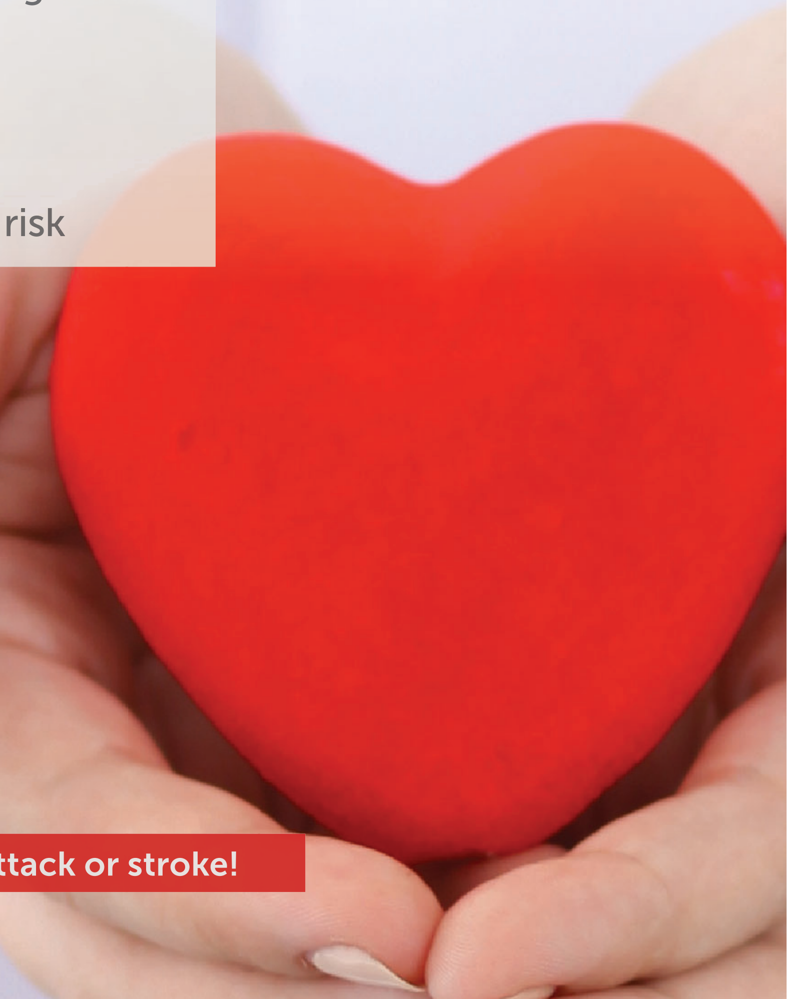

**Taking action now could prevent a heart attack or stroke!**

Hairdressers for

### Women's Health

- Regular health checks and screening can help to prevent illness and disease
- No matter how well you feel, there are important health recommendations to consider as you get older to help ensure optimal health and well-being

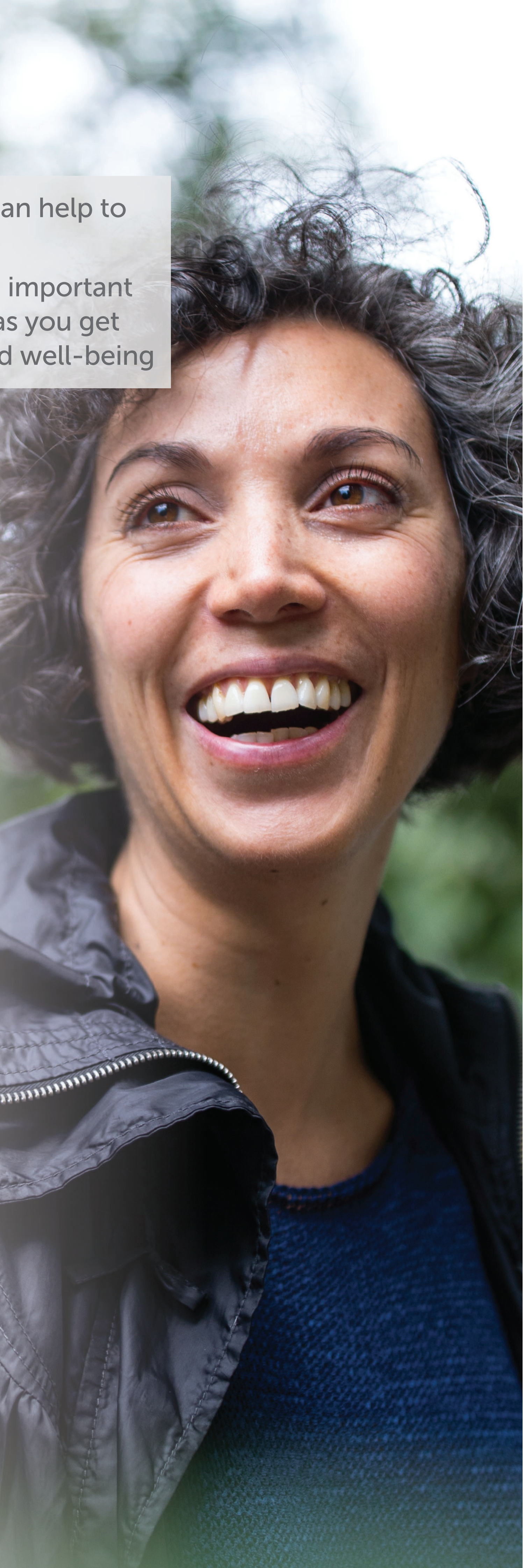

**Though these recommendations will vary for some women, for most women this includes:**

- ***CANCER SCREENING***
  Such as breast cancer screening at least every 2 years from the age 50

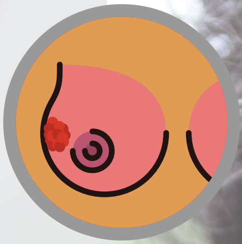

- ***EAT FOR HEALTH***
  Choose a variety of foods from each of the 5 food groups

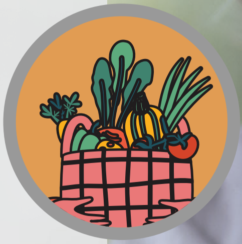

- ***SELF CARE***
  Make time to do the little things that nourish your soul.

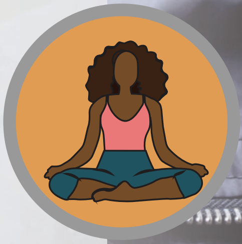

- ***STAY ACTIVE***
  Try for 150 or more minutes a week of physical activity

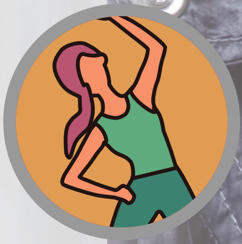

- ***REDUCE ALCOHOL***
  No more than 10 standard drinks a week

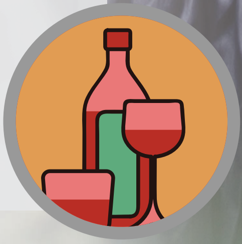

**Baseline Health Questionnaire - Hairdressers for Women's Health Study:**

**Registration data will include:**

Full Name

DOB

Mobile Ph no.

Email (with confirmation of email matching required)

GP Contact details

**Baseline Health Questionnaire:**

1. What is the highest level of education you have achieved?

A. Primary School
B. High School up to and including year 11
C. Year 12 High School
D. Tertiary Certificate

2. Where do you live in NSW?

*Please enter your 4 digit postcode*

3. What language do you mainly speak at home? (drop down list)

English
Mandarin
Arabic
Cantonese
Vietnamese
Italian
Greek
Hindi
Spanish
Punjabi
Tagalog
An Aboriginal language (please specify)
A Torres Strait Islander language (please specify)
Other (please specify)

4. In which country were you born? (drop down list)

5. Are you of Aboriginal or Torres Strait Islander Origin?

A. No
B. Yes, Aboriginal
C. Yes, Torres Strait Islander

D. Yes, both Aboriginal and Torres Strait Islander
E. Don't want to disclose

6. Have you been diagnosed with any of the following past or current health conditions by a doctor?

*Please click on Yes/No for each condition*

| ***Conditions*** | ***No*** | ***Yes*** |
|---|---|---|
| High blood pressure | | |
| High cholesterol | | |
| Asthma | | |
| Diabetes | | |
| Thyroid disorder | | |
| Chronic back pain | | |
| Rheumatoid arthritis | | |
| Inflammatory disease such as lupus (non Rheumatic) | | |
| Stomach problem such as an ulcer or gastritis or reflux | | |
| Colon problem such as irritable bowel or colitis | | |
| Poor blood circulation in your legs | | |
| Overweight | | |
| Hard of hearing | | |
| Vision problem | | |
| Chronic bronchitis or emphysema | | |
| Osteoarthritis ("regular" arthritis, not rheumatoid arthritis) | | |
| Osteoporosis (thinning of the bones) | | |
| Cancer during the last 5 years (not including small skin cancers) | | |
| Congestive heart failure | | |
| Heart disease such as angina (chest pain from heart problem), heart attack, bypass surgery or angioplasty | | |
| Stroke or TIA (transient ischaemic attack) | | |
| Kidney disease | | |
| Depression/ Anxiety | | |
| High blood pressure in Pregnancy | | |
| Diabetes in Pregnancy | | |
| Early birth of a child before 36 weeks gestation | | |
| Preeclampsia in pregnancy | | |
| Early Menopause before age 40years | | |

Do you have any other medical conditions that were not mentioned above? If so, please list them here:

7. Is there a family history of heart attacks or stroke before the age of 55 in a man or before the age of 65 in a woman in a first degree relative (mother, father, brother, sister)?
   Yes
   No

8. Is there a family history of cancer in a first degree relative (mother, father, brother, sister)?
   Yes
   No

9. Do you currently smoke cigarettes?
   Yes
   No

10. Have you ever smoked cigarettes (this includes those who have smoked > 100 cigarettes or a pipe/cigar or other form of tobacco > 20 times in your life?)
   Yes
   No

11. The following question is about your physical activity on an average week.
   Do you undertake physical activity for fitness, recreation or walking for transport that amounts to:

   At least 20 minutes of vigorous-intensity activity per day on at least 3 days per week (TOTAL = 60minutes/week)?
   *This refers to physical activity undertaken by adults for fitness, recreation, or sport that caused a respondent to breathe harder or puff and pant. This does not include walking, moderate physical activity, household chores, or vigorous gardening/yard work.*

   *Or*

   At least 30 minutes of moderate-intensity activity or walking per day on at least 5 days per week (TOTAL = 150minutes/week)?
   *Moderate = This refers to physical activity undertaken by adults for fitness, recreation, or sport that was more moderate, and not already reported as vigorous physical activity.*

YES
NO

12. How many serves of fruit do you eat on average per day?
    <2
    2-4
    >4

13. How many serves of Vegetables do you eat a day?
    <5
    5-7
    >7

14. EuroQOL 5D-5L Questionnaire
*Under each heading, please tick the ONE box that best describes your health TODAY*.

**MOBILITY**

I have no problems in walking about ☐

I have slight problems in walking about ☐

I have moderate problems in walking about ☐

I have severe problems in walking about ☐

I am unable to walk about ☐

**SELF-CARE**

I have no problems washing or dressing myself ☐

I have slight problems washing or dressing myself ☐

I have moderate problems washing or dressing myself ☐

I have severe problems washing or dressing myself ☐

I am unable to wash or dress myself ☐

**USUAL ACTIVITIES** (e.g. work, study, housework, family or leisure activities)

I have no problems doing my usual activities ☐

I have slight problems doing my usual activities ☐

I have moderate problems doing my usual activities ☐

I have severe problems doing my usual activities ☐

I am unable to do my usual activities ☐

**PAIN / DISCOMFORT**

I have no pain or discomfort ☐

I have slight pain or discomfort ☐

I have moderate pain or discomfort ☐

I have severe pain or discomfort ☐

I have extreme pain or discomfort ☐

**ANXIETY / DEPRESSION**

I am not anxious or depressed ☐

I am slightly anxious or depressed ☐

I am moderately anxious or depressed ☐

I am severely anxious or depressed ☐

I am extremely anxious or depressed ☐

We would like to know how good or bad your health is TODAY.

- This scale is numbered from 0 to 100.
- 100 means the best health you can imagine.
- 0 means the worst health you can imagine.
- Please mark an X on the scale to indicate how your health is TODAY.
- Now, write the number you marked on the scale in the box below

*(Please see scale overleaf)*

YOUR ☐ HEALTH TODAY =

Y.

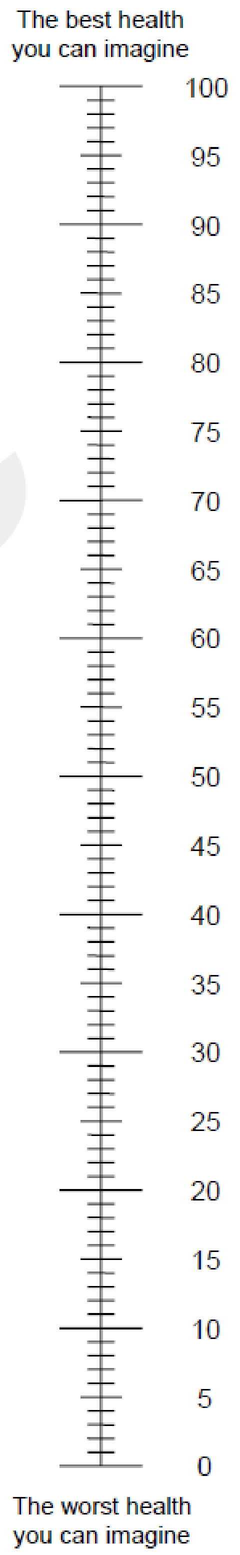
The best health
you can imagine
100
95
90
85
80
75
70
65
60
55
50
45
40
35
30
25
20
15
10
5
0
The worst health
you can imagine

The George Institute for Global Health

**6 month Health Questionnaire - Hairdressers for Women’s Health Study:**

**Registration data will include:**

Full Name

DOB

Mobile Ph no.

Email (with confirmation of email matching required)

GP contact details:

**6 month Health Questionnaire: The following questions were asked at the beginning of the study, we are now asking about the last 6 months.**

*15.* Have you been diagnosed with any of the following past or current health conditions by a doctor in the last 6 months?
*Please click on Yes/No for each condition*

| ***Conditions*** | ***No*** | ***Yes*** |
|---|---|---|
| High blood pressure | | |
| High cholesterol | | |
| Asthma | | |
| Diabetes | | |
| Thyroid disorder | | |
| Chronic back pain | | |
| Rheumatoid arthritis | | |
| Inflammatory disease such as lupus (non Rheumatic) | | |
| Stomach problem such as an ulcer or gastritis or reflux | | |
| Colon problem such as irritable bowel or colitis | | |
| Poor blood circulation in your legs | | |
| Overweight | | |
| Hard of hearing | | |
| Vision problem | | |
| Chronic bronchitis or emphysema | | |
| Osteoarthritis (“regular” arthritis, not rheumatoid arthritis) | | |
| Osteoporosis (thinning of the bones) | | |
| Cancer during the last 5 years (not including small skin cancers) | | |
| Congestive heart failure | | |

| |
|---|
| Heart disease such as angina (chest pain from heart problem), heart attack, bypass surgery or angioplasty |
| Stroke or TIA (transient ischaemic attack) |
| Kidney disease |
| Depression/ Anxiety |
| High blood pressure in Pregnancy |
| Diabetes in Pregnancy |
| Early birth of a child before 36 weeks gestation |
| Preeclampsia in pregnancy |
| Early Menopause before age 40years |

Do you have any other medical conditions diagnosed in the last 6 months that were not mentioned above? If so, please list them here:

16. Did you attend your General Practitioner in the 6 months since you commenced the study (virtually or face to face)?
    a. Yes because of the study (continue to Q3)
    b. Yes, for a reason other than this study
    c. No (skip to Q7)

17. When you attended your GP did you have a Heart Health Check as a result of being a part of this study?
    a. Yes (skip to Q7)
    b. No (continue to Q4)

18. Did you discuss a Heart Health check with your GP?
    a. Yes
    b. No

19. Why did you NOT have a heart health check? (please choose one response)
    a. GP decision
    b. Personal decision
    c. Joint decision
    d. COVID related issues
    e. Other (please describe)________________

20. Did your GP assess your heart risk by taking measurements or asking you related questions during a visit in the last 6 months? Please tick all that apply.

    a. blood pressure

    b. cholesterol

    c. sugar levels

    d. weight

    e. medical/family history

    f. physical activity

    g. diet

    h. smoking

21. Did your involvement in this study result in any health or behaviour related changes for you? For example changes in your exercise levels or diet or weight loss?
    a. Yes
    b. No

22. Did your involvement in this study result in any further investigations or additional medical appointments? For example specialist referral, or blood tests or scans?
    a. Yes (please comment)
    b. No

23. Is there a family history of heart attacks or stroke before the age of 55 in a man or before the age of 65 in a woman in a first degree relative (mother, father, brother, sister)?
    Yes
    No

24. Is there a family history of cancer in a first degree relative (mother, father, brother, sister)?
    Yes
    No

25. Do you currently smoke cigarettes?
    Yes
    No

26. Have you ever smoked cigarettes (this includes those who have smoked > 100 cigarettes or a pipe/cigar or other form of tobacco > 20 times in your life?)
    Yes
    No

27. The following question is about your physical activity on an average week.
    Do you undertake physical activity for fitness, recreation or walking for transport that amounts to:

    At least 20 minutes of vigorous-intensity activity per day on at least 3 days per week (TOTAL = 60minutes/week)?
    *This refers to physical activity undertaken by adults for fitness, recreation, or sport that caused a respondent to breathe harder or puff and pant. This does not include walking, moderate physical activity, household chores, or vigorous gardening/yard work.*

    *Or*

    At least 30 minutes of moderate-intensity activity or walking per day on at least 5 days per week (TOTAL = 150minutes/week)?
    *Moderate = This refers to physical activity undertaken by adults for fitness, recreation, or sport that was more moderate, and not already reported as vigorous physical activity.*

    YES
    NO

28. How many serves of fruit do you eat on average per day?
    <2
    2-4
    >4

29. How many serves of Vegetables do you eat a day?
    <5
    5-7
    >7

30. EuroQOL 5D-5L Questionnaire

*Under each heading, please tick the ONE box that best describes your health TODAY*.

**MOBILITY**

I have no problems in walking about ☐

I have slight problems in walking about ☐

I have moderate problems in walking about ☐

I have severe problems in walking about ☐

I am unable to walk about ☐

**SELF-CARE**

I have no problems washing or dressing myself ☐

I have slight problems washing or dressing myself ☐

I have moderate problems washing or dressing myself ☐

I have severe problems washing or dressing myself ☐

I am unable to wash or dress myself ☐

**USUAL ACTIVITIES** (e.g. work, study, housework, family or leisure activities)

I have no problems doing my usual activities ☐

I have slight problems doing my usual activities ☐

I have moderate problems doing my usual activities ☐

I have severe problems doing my usual activities ☐

I am unable to do my usual activities ☐

**PAIN / DISCOMFORT**

I have no pain or discomfort ☐

I have slight pain or discomfort ☐

I have moderate pain or discomfort ☐

I have severe pain or discomfort ☐

I have extreme pain or discomfort ☐

**ANXIETY / DEPRESSION**

I am not anxious or depressed ☐

I am slightly anxious or depressed ☐

I am moderately anxious or depressed ☐

I am severely anxious or depressed ☐

I am extremely anxious or depressed ☐

We would like to know how good or bad your health is TODAY.

- This scale is numbered from 0 to 100.
- 100 means the best health you can imagine.
- 0 means the worst health you can imagine.
- Please mark an X on the scale to indicate how your health is TODAY.
- Now, write the number you marked on the scale in the box below

*(Please see scale overleaf)*

YOUR ☐ HEALTH TODAY =

Y.

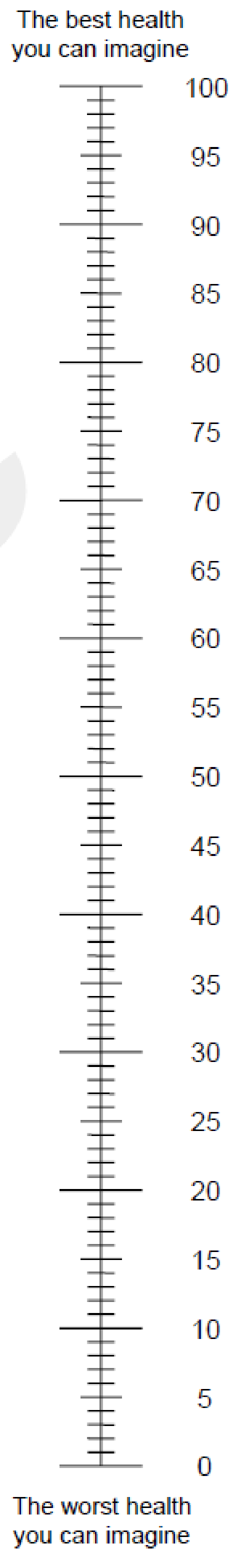
The best health you can imagine
100
95
90
85
80
75
70
65
60
55
50
45
40
35
30
25
20
15
10
5
0
The worst health you can imagine

**RECORD OF INTERVIEW FOR CLIENTS FOR THE HAIRDRESSERS FOR HEART HEALTH STUDY.**

**Name of Interviewer:** ______________________________________________

**Date**: __ __/ __ __ /__ __ __ __
d d / m m/ y y y y

**PE ID No:** ______________________________________________

**Post code:**

**Date of birth**: __ __/ __ __ /__ __ __ __
d d / m m/ y y y y

**RESEARCHER FIELD NOTES:**

How do you think the interview went?

What struck you as important?

What further questions/areas would you like to explore in the next interview?

**INTERVIEW GUIDE FOR CONSUMERS**

**Overall aim of the process evaluation interviews:** To understand for whom, and how and why 'nudging' women through sharing of educational information, and a letter to the GP, at their hair dresser salons, worked(or not) in following up with their general practitioners to get a heart health check done.

**Say to the consumer**

*Hi, I am [name] and I'm from [organisation]. Thank you for taking part in this interview. As discussed, we are trying to find out how we can better care for women with heart disease. You can change your mind about talking to me at any time before or during the interview and stop the interview at any time. Are you happy to continue? [If no, thank them for their time and end interview; if yes continue.] Thank you [name] for agreeing to take part.*

*We are interviewing women who have been involved in the study. Are you happy for me to record the interview? Our interview will be transcribed professionally and we will ensure your privacy and confidentiality.*

*Do you have any questions before we start?*

*Note: Key questions in bold, with probing questions in non-bold. Questions do not have to be asked in this order, and not all questions have to be covered.

| |
|---|
| **<u>CONTEXT DOMAIN: To understand the context of the women managing their cardiovascular risk and how hairdressers for heart health project fits within this</u>** |
| **What were your first impressions about the <u>Hairdressers for heart health study?</u> What do you think the study would like to achieve**? [probe: their knowledge, attitudes and behaviour of CVD risk factors- smoking, hypertension, exercise, diet]<br><br>**What do you know about cardiovascular disease?** [probe: what they know about it, e.g. family history, risk factors, public health campaigns, gender differences,<br><br>**What do you know about your own cardiovascular risk, where do you assess such information?** [probe: use of other google searches, services/platforms/apps, trusted health providers]<br><br>**Have you seen the GP in the last 6 months? What was the visit for?** [probe: their views on prevention, women's health] |
| **<u>IMPLEMENTATION DOMAIN: To understand the acceptability, adoption, implementation barriers and facilitators</u>** |
| **Can you tell me more about your thoughts in taking part in the study? What was good or bad about it? [**probe: initial contact at the hairdressers, thoughts about the materials for both usual care and intervention arm]<br><br>**FOR INTERVENTION ARM:**<br>• **What did you think about the materials that were given to you at the hairdressers?** [probe: new knowledge, relevance to their daily life]<br><br>• **Did you have a heart health check done? Why or why not? What did you think about it?** [probe: any changes in management, life-style factors etc]<br><br>• **Did the letter provided play a part in supporting your decision to see your GP? Why or why not? [**probe: competing priorities,]<br><br>**What do you think about this approach, will it be useful in reaching women who have higher cardiovascular risk, to have a heart health check?** [probe: is hairdressers salon a good setting, privacy issues]<br><br>**Were there any external issues that impacted upon your thoughts about the project? [**probes: impact of COVID in seeing GPs] |
| **<u>MECHANISMS DOMAIN: To understand the outcomes and future implementation</u>** |
| **Upon reflection, do you think this study will increase the awareness, and screening of women for their cardiovascular risk? Why or why not? Can you elaborate on some of the reasons?** [probe: patient factors, hairdresser, general community awareness factors]<br>• Upon hindsight, is there anything that we should do differently?<br><br>**Would it be beneficial and feasible to continue this community outreach to women through their hairdressers or other settings?**<br>• Do you think this will be widely accepted by women? Why or why not? |
| **Concluding question and Statement** |
| Is there anything else you would like to say that we have not talked about in this interview?<br><br>Thank you so much for your time and for sharing your insights. |

**RECORD OF INTERVIEW FOR HAIRDRESSER SALON STAFF FOR THE HAIRDRESSERS FOR HEART HEALTH STUDY.**

**Name of Interviewer:** ____________________________________________

**Date**: __ __/ __ __ /__ __ __ __
d d / m m / y y y y

**PE ID No:** ____________________________________________

**Place of employment:** ____________________________________________

**Date of birth**: __ __/ __ __ /__ __ __ __
d d / m m / y y y y

**Gender**: Female / Male / Non Binary

**RESEARCHER FIELD NOTES:**

How do you think the interview went?

What struck you as important?

What further questions/areas would you like to explore in the next interview?

**INTERVIEW GUIDE FOR HAIRDRESSERS**

**Overall aim of the process evaluation interviews:** To understand for whom, and how and why 'nudging' women through sharing of educational information, and a letter to the GP, at their hair dresser salons, worked(or not) in following up with their general practitioners to get a heart health check done.

**Say to the hairdresser staff:**

*Hi, I am [name] and I'm from [organisation]. Thank you for taking part in this interview. As discussed, we are trying to find out how we can better care for women with heart disease. You can change your mind about talking to me at any time before or during the interview and stop the interview at any time. Are you happy to continue? [If no, thank them for their time and end interview; if yes continue.] Thank you [name] for agreeing to take part.*

*We are interviewing hairdressers who have been involved in the study. Are you happy for me to record the interview? Our interview will be transcribed professionally and we will ensure your privacy and confidentiality.*

*Do you have any questions before we start?*

*Note: Key questions in bold, with probing questions in non-bold. Questions do not have to be asked in this order, and not all questions have to be covered.

| **<u>CONTEXT DOMAIN: To understand the context of the hairdresser salons, and how hairdressers for heart health project fits within this</u>** |
|---|
| **Warm up: What is your role in this study? [**Probe: hairdresser, manager of the salon, related tasks**]**<br><br>**What were your first impressions about the <u>Hairdressers for heart health study?</u> What do you think the study would like to achieve**? [probe: their understanding of CVD risk, clients, impact on their work, interactions with their clients, time spent on the training, their thoughts about the materials]<br><br>**What are your thoughts about the study at your salon? What was good or bad about it? Were you comfortable providing the materials to your clients?** [probe: specifically the use of a non-health setting for health education, number and process of identifying suitable clients, underlying relationship/dynamic with clients, and providing project materials to your clients] |
| **<u>IMPLEMENTATION DOMAIN: To understand reach, acceptability, adoption, implementation barriers and facilitators</u>** |
| **What in your perspective has gone well in this project? Can you provide some examples.** [probe: approaching all suitable clients, clients' reactions, training, reimbursement, keeping logs or data, logistics e.g. print outs etc]<br><br>**What in your perspective did you not like about this project? Why is that?** [probe: clients' reactions, time taken, impact on the salon.]<br><br>**What do you think of the adoption of this approach, will it be useful in reaching women who may have higher cardiovascular risk, in the long term?** [probe: is hairdressers salon a good setting, or other settings]<br><br>**Were there any system issues that impacted upon the implementation?** [probe about: workforce, reimbursement, impact of COVID] |
| **<u>MECHANISMS DOMAIN: To understand the mechanisms, that would impact on the outcomes and future implementation</u>** |
| **Do you think this study will increase the awareness, and screening of women for their cardiovascular risk? Why or why not? Can you elaborate on some of the reasons?** [probe: patient factors, hairdresser, general community awareness factors]<br>• Upon hindsight, is there anything that we should do differently?<br><br>**Would it be beneficial and feasible to continue this community outreach to women through their hairdressers or other settings?**<br>• Do you think this will be widely accepted by clients and other hairdressers? Why or why not? |
| **Concluding question and Statement** |
| Is there anything else you would like to say that we have not talked about in this interview?<br><br>Thank you so much for your time and for sharing your insights. |

**Appendix. Standards for reporting qualitative research (SRQR) Checklist**

| Topic | | Page # |
|---|---|---|
| **Title and abstract** | | |
| | **Title** - Concise description of the nature and topic of the study Identifying the study as qualitative or indicating the approach (e.g., ethnography, grounded theory) or data collection methods (e.g., interview, focus group) is recommended | 1 |
| | **Abstract** - Summary of key elements of the study using the abstract format of the intended publication; typically includes background, purpose, methods, results, and conclusions | 2 |
| **Introduction** | | |
| | **Problem formulation** - Description and significance of the problem/phenomenon studied; review of relevant theory and empirical work; problem statement | 4-5 |
| | **Purpose or research questio**n - Purpose of the study and specific objectives or questions | 5 |
| **Methods** | | |
| | **Qualitative approach and research paradigm** - Qualitative approach (e.g., ethnography, grounded theory, case study, phenomenology, narrative research) and guiding theory if appropriate; identifying the research paradigm (e.g., postpositivist, constructivist/ interpretivist) is also recommended; rationale** | 7-9 |
| | **Researcher characteristics and reflexivity** - Researchers' characteristics that may influence the research, including personal attributes, qualifications/experience, relationship with participants, assumptions, and/or presuppositions; potential or actual interaction between researchers' characteristics and the research questions, approach, methods, results, and/or transferability | 9 |
| | **Context** - Setting/site and salient contextual factors; rationale** | 8 |
| | **Sampling strategy** - How and why research participants, documents, or events were selected; criteria for deciding when no further sampling was necessary (e.g., sampling saturation); rationale** | 8- 9 |
| | **Ethical issues pertaining to human subjects** - Documentation of approval by an appropriate ethics review board and participant consent, or explanation for lack thereof; other confidentiality and data security issues | 6 |
| | **Data collection methods** - Types of data collected; details of data collection procedures including (as appropriate) start and stop dates of data collection and analysis, iterative process, triangulation of sources/methods, and modification of procedures in response to evolving study findings; rationale** | 8-9 |
| | **Data collection instruments and technologies** - Description of instruments (e.g., interview guides, questionnaires) and devices (e.g., audio recorders) used for data collection; if/how the instrument(s) changed over the course of the study | 9 |

| | | |
|---|---|---|
| | **Units of study** - Number and relevant characteristics of participants, documents, or events included in the study; level of participation (could be reported in results) | 9 |
| | **Data processing** - Methods for processing data prior to and during analysis, including transcription, data entry, data management and security, verification of data integrity, data coding, and anonymization/de-identification of excerpts | 9 |
| | **Data analysis** - Process by which inferences, themes, etc., were identified and developed, including the researchers involved in data analysis; usually references a specific paradigm or approach; rationale** | 9 |
| | **Techniques to enhance trustworthiness** - Techniques to enhance trustworthiness and credibility of data analysis (e.g., member checking, audit trail, triangulation); rationale** | 9 |
| **Results/findings** | | |
| | **Synthesis and interpretation** - Main findings (e.g., interpretations, inferences, and themes); might include development of a theory or model, or integration with prior research or theory | 9-13 |
| | **Links to empirical data** - Evidence (e.g., quotes, field notes, text excerpts, photographs) to substantiate analytic findings | 9-13 |
| **Discussion** | | |
| | **Integration with prior work, implications, transferability, and contribution(s) to the field -** Short summary of main findings; explanation of how findings and conclusions connect to, support, elaborate on, or challenge conclusions of earlier scholarship; discussion of scope of application/generalizability; identification of unique contribution(s) to scholarship in a discipline or field | 13-17 |
| | **Limitations** - Trustworthiness and limitations of findings | 16-17 |
| **Other** | | |
| | **Conflicts of interest** - Potential sources of influence or perceived influence on study conduct and conclusions; how these were managed | 18 |
| | **Funding** - Sources of funding and other support; role of funders in data collection, interpretation, and reporting | 19 |
| | *The authors created the SRQR by searching the literature to identify guidelines, reporting standards, and critical appraisal criteria for qualitative research; reviewing the reference lists of retrieved sources; and contacting experts to gain feedback. The SRQR aims to improve the transparency of all aspects of qualitative research by providing clear standards for reporting qualitative research. | |
| | **The rationale should briefly discuss the justification for choosing that theory, approach, method, or technique rather than other options available, the assumptions and limitations implicit in those choices, and how those choices influence study conclusions and transferability. As appropriate, the rationale for several items might be discussed together. | |
| | **Reference:** | |

| | |
|---|---|
| | O'Brien BC, Harris IB, Beckman TJ, Reed DA, Cook DA. **Standards for reporting qualitative research: a synthesis of recommendations.** *Academic Medicine*, Vol. 89, No. 9 / Sept 2014 DOI: 10.1097/ACM.0000000000000388 |
